# Patterns and Predictors of Artificial Intelligence Use Among Healthcare Professionals in the United States and United Kingdom: A Cross-National Survey

**DOI:** 10.64898/2026.05.01.26352171

**Authors:** Emre Sezgin, Jennifer A. Lee, Tomasz Jadczyk, Alysha J Taxter

## Abstract

**Objective:** We surveyed 524 healthcare professionals (HCPs) in the United States and United Kingdom to examine workplace generative AI use, access, and barriers in two high-maturity health settings.

**Methods:** This cross-sectional survey compared AI usage breadth, access modes, and barriers among HCPs, stratified by country and professional role.

**Results:** Overall, 75.8% of HCPs reported recent AI use, mainly for documentation, literature search, and clinical decision support. Usage breadth was similar by country, but role differences were pronounced. Physicians reported broader use and were significantly more likely to access AI via personal, non-employer-provided tools (60.4% vs. 31.0% for nurses; P<.01). Personal tools were the most common access mode overall (40.1%).

**Conclusion:** AI use is common, but institutional access lags adoption. Shifting use from personal accounts toward governed, approved systems is a key priority.

## Introduction

Generative artificial intelligence (AI) is increasingly entering clinical work, but evidence remains limited on how health care professionals (HCPs) use AI in routine practice and whether use occurs within sanctioned systems or outside institutional governance.^1^ Because the United Kingdom (UK) and United States (US) differ in health system organization and are both high-maturity AI-in-health settings,^2^ we examined workplace AI use, access routes, and barriers among HCPs in both countries.

## Methods

We conducted a cross-sectional online survey of HCPs working in medicine in the UK and US. The primary exposure was country; secondary analyses compared physicians, nurses, and other HCPs. AI usage breadth was defined as the number of AI application types used for work (range, 0-9). Categorical comparisons used χ^2^ or Fisher exact tests, and univariate linear regression examined breadth. Analyses were performed in Stata 18.0. The Nationwide Children’s Hospital institutional review board approved the study (IRB #00005734), and we followed the STROBE reporting guidelines. See Supplementary Materials S1 for details on study methodology.

## Results

Among 524 respondents, 268 (51.1%) were from the UK and 256 (48.9%) from the US; 281 (53.6%) were nurses, 111 (21.2%) physicians, and 132 (25.2%) other HCPs. Most were female (386 [73.7%]) and employed full-time (390 [74.4%]); country groups differed in education and experience (p<0.05) but not age or specialty.(See Supplementary Material S2 for descriptive statistics table)

Overall, 397 respondents (75.8%) reported at least one workplace AI use in the prior 30 days. The most common uses were documentation support (191 [36.5%]), literature search/summarization (158 [30.2%]), and clinical decision support (CDS; 128 [24.4%]) (Table 1). Any nonuse did not differ by country (UK, 24.3%; US, 24.2%; P=.99), and AI usage breadth was similar (B=0.19; 95% CI, -0.04 to 0.42; P=.11). However, US respondents more often reported CDS use (28.9% vs 20.1%; P=.02), imaging/diagnostics support (10.5% vs 6.0%; P=.05), and coding/billing/prior authorization use (9.0% vs 3.4%; P<.01), whereas UK respondents more often reported translation/language support (12.7% vs 4.7%; P<.01).

**Table 1.** AI Tool Use in the Past 30 Days, by Country of Residence and Professional Role.

| AI tool type used in past 30 days | Country of Residence |  |  | Professional Role |  |  | p-value (role) |
| --- | --- | --- | --- | --- | --- | --- | --- |
|  | Total(N=524) | United Kingdom(N=268) | United States(N=256) | Physician(N=111) | Nurse(N=281) | Other(N=132) |  |
| Documentation | 191 (36.5%) | 90 (33.6%) | 101 (39.5%) | 50 (45.0%) | 98 (34.9%) | 43 (32.6%) | 0.10 |
| Clinical decision support | 128 (24.4%) | 54 (20.1%) | 74 (28.9%) | 49 (44.1%) | 62 (22.1%) | 17 (12.9%) | <0.01 |
| Literature search / summarization | 158 (30.2%) | 79 (29.5%) | 79 (30.9%) | 47 (42.3%) | 65 (23.1%) | 46 (34.8%) | <0.01 |
| Data analysis / quality improvement | 84 (16.0%) | 39 (14.6%) | 45 (17.6%) | 17 (15.3%) | 40 (14.2%) | 27 (20.5%) | 0.27 |
| Patient messaging drafts | 69 (13.2%) | 34 (12.7%) | 35 (13.7%) | 19 (17.1%) | 32 (11.4%) | 18 (13.6%) | 0.31 |
| Imaging / diagnostics support | 43 (8.2%) | 16 (6.0%) | 27 (10.5%) | 16 (14.4%) | 24 (8.5%) | 2 (1.5%) | <0.01 |
| Translation / language support | 46 (8.8%) | 34 (12.7%) | 12 (4.7%) | 9 (8.1%) | 25 (8.9%) | 12 (9.1%) | 0.96 |
| Coding / billing / prior authorization | 32 (6.1%) | 9 (3.4%) | 23 (9.0%) | 10 (9.0%) | 13 (4.6%) | 9 (6.8%) | 0.24 |
| Other AI use / not listed | 41 (7.8%) | 25 (9.3%) | 16 (6.3%) | 5 (4.5%) | 27 (9.6%) | 9 (6.8%) | 0.21 |
| Not used - any AI tools | 127 (24.2%) | 65 (24.3%) | 62 (24.2%) | 17 (15.3%) | 81 (28.8%) | 29 (22.0%) | 0.02 |

*Results are not mutually exclusive (respondents could select multiple tool types). Country p-values are from chi-square tests. Country differences were significant for clinical decision support ( $p=0.02$ ) and translation/language support ( $p<0.01$ ); all other country comparisons, including other AI use/not listed, had $p\geq 0.08$ .*

Role differences were larger than country differences. Physicians more often reported CDS use than nurses and other HCPs (44.1% vs 22.1% vs 12.9%; P<.01), as well as greater use of literature search and imaging/diagnostics tools (both P<.01). Physicians were also less likely to report no AI use (15.3% vs 28.8% among nurses and 22.0% among other HCPs; P=.02). In univariate models, nurse and other HCP roles were associated with lower AI breadth than physician role; graduate or doctoral education, male sex, and full-time employment were associated with greater breadth (See Supplementary Material S3 for regression analysis results).

No single barrier predominated (Table 2). The most common barriers were accuracy or reliability concerns (78 [14.9%]), lack of workplace access (77 [14.7%]), privacy or security concerns (65 [12.4%]), and lack of training (63 [12.0%]); barrier distributions did not differ significantly by country (P=.055). Personal, non-employer-provided tools were the most common access mode (210 [40.1%]), exceeding EHR-integrated access (128 [24.4%]) and employer-approved standalone tools (134 [25.6%]). Physicians were more likely than nurses to report personal tool use (60.4% vs 31.0%; P<.01), whereas EHR-integrated and employer-approved access did not differ by role. The most commonly endorsed facilitators were stronger evidence (227 [43.3%]), organizational policy (156 [29.8%], training (155 [29.6%]), privacy protections (149 [28.4%]), and EHR integration (130 [24.8%]).

**Table 2.**
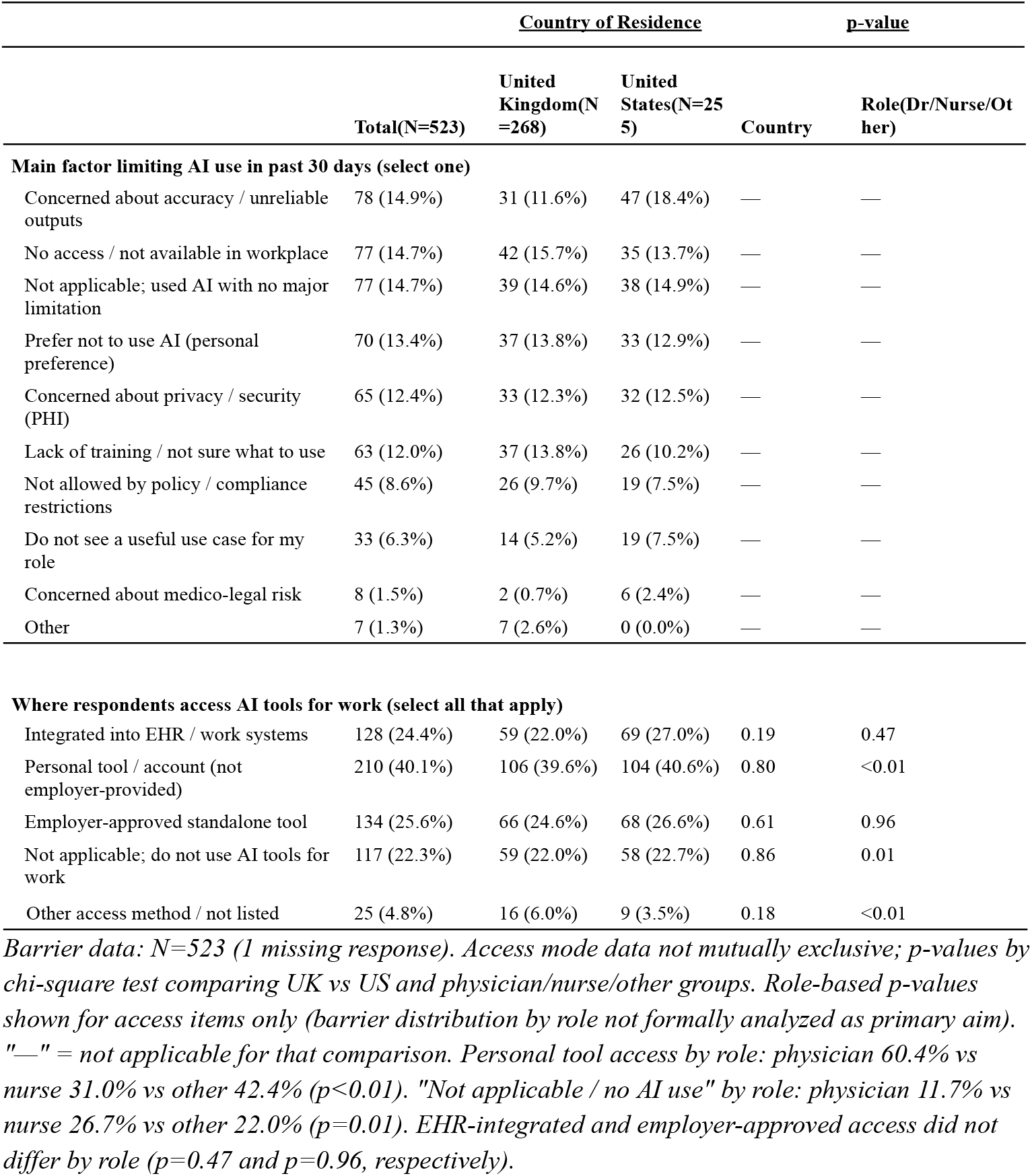
Barriers to AI Use and AI Access Mode, Overall, by Country of Residence, and by Professional Role.

|  | Country of Residence | p-value |
| --- | --- | --- |

|  | Total(N=523) | United Kingdom(N=268) | United States(N=255) | Country | Role(Dr/Nurse/Other) |
| --- | --- | --- | --- | --- | --- |
| **Main factor limiting AI use in past 30 days (select one)** |  |  |  |  |  |
| Concerned about accuracy / unreliable outputs | 78 (14.9%) | 31 (11.6%) | 47 (18.4%) | — | — |
| No access / not available in workplace | 77 (14.7%) | 42 (15.7%) | 35 (13.7%) | — | — |
| Not applicable; used AI with no major limitation | 77 (14.7%) | 39 (14.6%) | 38 (14.9%) | — | — |
| Prefer not to use AI (personal preference) | 70 (13.4%) | 37 (13.8%) | 33 (12.9%) | — | — |
| Concerned about privacy / security (PHI) | 65 (12.4%) | 33 (12.3%) | 32 (12.5%) | — | — |
| Lack of training / not sure what to use | 63 (12.0%) | 37 (13.8%) | 26 (10.2%) | — | — |
| Not allowed by policy / compliance restrictions | 45 (8.6%) | 26 (9.7%) | 19 (7.5%) | — | — |
| Do not see a useful use case for my role | 33 (6.3%) | 14 (5.2%) | 19 (7.5%) | — | — |
| Concerned about medico-legal risk | 8 (1.5%) | 2 (0.7%) | 6 (2.4%) | — | — |
| Other | 7 (1.3%) | 7 (2.6%) | 0 (0.0%) | — | — |

**Where respondents access AI tools for work (select all that apply)**
|  |  |  |  |  |  |
| --- | --- | --- | --- | --- | --- |
| Integrated into EHR / work systems | 128 (24.4%) | 59 (22.0%) | 69 (27.0%) | 0.19 | 0.47 |
| Personal tool / account (not employer-provided) | 210 (40.1%) | 106 (39.6%) | 104 (40.6%) | 0.80 | <0.01 |
| Employer-approved standalone tool | 134 (25.6%) | 66 (24.6%) | 68 (26.6%) | 0.61 | 0.96 |
| Not applicable; do not use AI tools for work | 117 (22.3%) | 59 (22.0%) | 58 (22.7%) | 0.86 | 0.01 |
| Other access method / not listed | 25 (4.8%) | 16 (6.0%) | 9 (3.5%) | 0.18 | <0.01 |

*Barrier data: N=523 (1 missing response). Access mode data not mutually exclusive; p-values by chi-square test comparing UK vs US and physician/nurse/other groups. Role-based p-values shown for access items only (barrier distribution by role not formally analyzed as primary aim). "—" = not applicable for that comparison. Personal tool access by role: physician 60.4% vs nurse 31.0% vs other 42.4% ( $p<0.01$ ). "Not applicable / no AI use" by role: physician 11.7% vs nurse 26.7% vs other 22.0% ( $p=0.01$ ). EHR-integrated and employer-approved access did not differ by role ( $p=0.47$ and $p=0.96$ , respectively).*

## Discussion

In this cross-national survey, three-quarters of respondents reported recent workplace AI use. Use was concentrated in documentation, information retrieval, and clinical decision support rather than imaging or revenue-cycle tasks, suggesting earlier adoption in activities where generative tools can be layered onto existing workflows with low implementation burden.^3,4^ Despite differences between the UK and US, overall adoption and usage breadth were similar, although the US more often reported clinical decision support, imaging, and coding/billing uses, whereas the UK more often reported translation support.^5^

Role-based differences were more pronounced. Physicians reported broader use and were substantially more likely to access AI through personal, non-employer-provided accounts. This pattern suggests both uneven workflow integration and a governance gap when clinician demand outpaces approved organizational access.

The barriers do not support a single implementation solution. Concerns about reliability, access, privacy, and training were all common, while medico-legal risk was infrequently identified as the limitation.^6^ This contrasts the literature that emphasizes tool maturity, cost, regulatory uncertainty, reimbursement, real-world evidence, and ethical/medicolegal governance.^3^ In the UK, the governance landscape itself is heterogeneous. NHS organizations operate under General Data Protection Regulation (GDPR) constraints that restrict the transfer of patient data to external systems but rely largely on individual professional responsibility rather than formal monitoring,^7^ whereas some private settings may impose stricter organizational restrictions on AI use for clinical documentation. Thus, “no access” may reflect appropriate avoidance of unsanctioned tools, organizational prohibition, or unmet demand (each requiring different implementation responses) and strategies should combine approved access, policy, privacy safeguards, evidence, and training.^8^

This study has limitations, including online panel recruitment, self-reported 30-day behavior, heterogeneity within the “other HCP” group, and a cross-sectional design that cannot establish whether access drives use or whether motivated users seek it out. Overall, workplace AI use was common, but institutional access lagged behind adoption;^9^ closing role-based gaps while shifting use from personal tools toward governed systems may be a practical near-term priority.^7,8^

## Data Availability

All data produced are available online at https://www.kaggle.com/datasets/IFRL-AWRI/usa-and-uk-healthcare-provider-survey-on-ai-use

## Dataset

Study dataset will be available at:

https://www.kaggle.com/datasets/IFRL-AWRI/usa-and-uk-healthcare-provider-survey-on-ai-use

## Supplementary Materials

### S1. Methods

This supplement provides additional methodological detail to support reproducibility and to document variable definitions used in the main manuscript.

#### Study design and recruitment

This study was an anonymous, cross-sectional online survey of healthcare professionals residing in the United Kingdom and United States. Participants were recruited through Prolific using prespecified eligibility criteria based on country of residence and current work in a healthcare profession. Eligible respondents were adults aged 18 years or older who reported current work in medicine or allied health roles. Participation was voluntary, and respondents reviewed electronic study information before survey initiation. No protected health information was collected. Survey responses were collected between January-March 2026.

#### Survey content

The questionnaire included items on country of residence, gender, sex, age group, educational attainment, years of work experience, employment status, professional role, primary clinical specialty, and whether English was the respondent’s primary language. Work-related AI use was assessed with the item, “In the past 30 days, which types of AI have you used for work?” (Table 1). Respondents could select multiple categories: documentation support, clinical decision support, literature search or summarization, data analysis or quality improvement, imaging or diagnostics support, patient messaging drafts, coding or billing or prior authorization help, translation or language support, other AI use, and “I have not used AI tools.” Barriers were assessed with a single-choice item asking for the main factor limiting AI use at work. Access routes and factors that would increase workplace AI use were assessed as multiple-response items.

**Table 1.** Variables and descriptions.

| Variable | Description | Type | Sample Values |
| --- | --- | --- | --- |
| Gender | The gender identity of the respondent. | Categorical | Woman (including Trans Female/Trans Woman), Man (including Trans Male/Trans Man) |
| Highest education level completed | The highest level of education attained by the respondent. | Ordinal/Categorical | Doctorate degree (PhD/other), Graduate degree (MA/MSc/MPhil/other), Undergraduate degree (BA/BSc/other), Technical/community college, High school diploma/A-levels |
| Years of work experience | The number of years the respondent has been working. | Ordinal/Categorical | 16+ years, 11-15 years, 6-10 years, 3-5 years, 1-2 years |
| Employment sector within medicine/healthcare | The respondent's employment sector within medicine/healthcare. | Categorical | Nurse, Doctor, Pharmacist, Psychologist, Social Worker |
| Age | The age of the respondent. | Numerical (Discrete) | Age range > 18 |
| Sex | The biological sex of the respondent. | Categorical | Female, Male |
| Ethnicity | A simplified categorization of the respondent's ethnicity. | Categorical | White, Asian, Black, Mixed, Other |
| Country of birth | The country where the respondent was born. | Categorical | United States, United Kingdom, Turkey, Nepal, Nigeria, Hong Kong, Portugal, Germany, Taiwan, Switzerland, Philippines |
| Country of residence | The country where the respondent currently resides. | Categorical | United States, United Kingdom |
| Nationality | The nationality of the respondent. | Categorical | United States, United Kingdom, Nepal, Nigeria, Germany, Hong Kong, Portugal, Taiwan |
| Language | The primary language of the respondent. | Categorical | English, Nepali, German, Cantonese, Portuguese, Tagalog-Filipino, Bengali |
| Employment status | The respondent's current employment status. | Categorical | Full-Time, Part-Time |
| What is your primary clinical specialty/area? | The respondent's main clinical specialty or area of practice. | Categorical | Primary care / general practice, Internal medicine subspecialty, Behavioral and mental health, Pharmacy, Emergency / urgent care, Care coordination and social services, Surgery (all surgical specialties), Pediatrics |
| In the past 30 days, which types of AI have you used for work? (Select all that apply) | Types of AI tools used for work in the past 30 days (multi-select). | Multi-select Categorical | Documentation (AI scribes/ambient note drafting; summaries; after-visit instructions), Clinical decision support (risk scores; alerts; recommendations), Literature search/ summarization, I have not used AI tools for work in the past 30 days, Data analysis / quality improvement, Imaging/ diagnostics support, Patient messaging drafts, Coding/ billing/ prior authorization help, Translation/ language support, Other AI use/ Not listed |
| In the past 30 days, what has been the main factor | The single main factor limiting AI use at work in the past 30 days. | Categorical | No access / not available in my workplace, Concerned about accuracy/unreliable outputs, Lack of training / Not sure what to use, Prefer not to use AI (personal preference), Concerned about privacy/security (PHI), Not allowed by policy / |
| LIMITING your use of AI tools at work? (Select one) |  |  | compliance restrictions, Do not see a useful use case for my role, Concerned about medico-legal risk, Not applicable; I used AI tools for work in the past 30 days and had no major limitations, Other reason/ not listed |
| What would make you MORE LIKELY to use AI at work? (Select up to 2) | Factors that would increase the likelihood of AI use at work (multi-select). | Multi-select Categorical | Proof of accuracy/safety for my tasks, Time savings demonstrated, Clear organizational policy, Education and training, Strong privacy safeguards, Electronic Medical Records (EMR) integration, Peer/leadership endorsement, Nothing; I have no interest, Nothing; I already use AI as much as I want, Other factor/ not listed |
| Where do you access AI tools for work? (Select all that apply) | The location or method of accessing AI tools for work (multi-select). | Multi-select Categorical | Integrated into EHR/work systems, Personal tool/account (not employer-provided), Employer-approved standalone tool, Not applicable; I do not use AI tools for work, Other access method / not listed |

#### Derived variables

Country of residence was the primary exposure. For role-stratified analyses, respondents were grouped as physician, nurse, or other healthcare professional. The “other” category included pharmacists, psychologists, social workers, and other nonphysician, nonnursing roles. The AI usage breadth score was defined as the count of positively endorsed work-related AI application types across 9 categories; the “I have not used AI tools” response was excluded from score construction. Possible scores therefore ranged from 0 to 9. Because AI-use, access, and facilitator items were multiple-response questions, category percentages were not mutually exclusive. (Table 2)

**Table 2.**
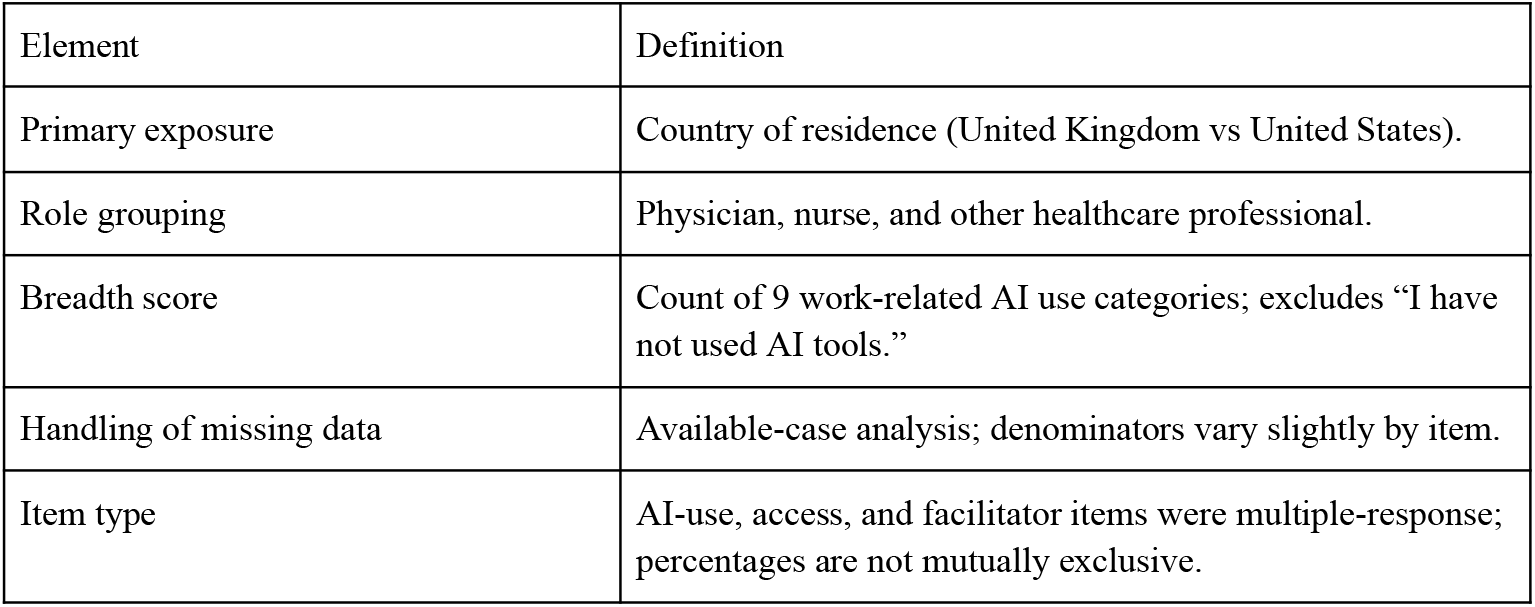
Core analytic definitions.

#### Statistical analysis

Analyses were descriptive and exploratory. Categorical variables were summarized as counts and percentages. Associations for categorical comparisons were evaluated with chi-square tests; Fisher exact tests were used when expected cell counts were sparse. Associations with the AI usage breadth score were evaluated using separate univariate linear regression models, reported as B coefficients with 95% confidence intervals. Two-sided P values less than .05 were interpreted as statistically significant.

Analyses were conducted using Stata version 18.0.

#### Missing data and denominators

Missing item responses were uncommon and were handled by available-case analysis. As a result, denominators varied slightly across analyses, with most items using N=524 and selected items using N=523 because of 1 missing response. Table footnotes in the main manuscript report when results are based on multiple-response items and when denominators differ across variables.

#### Data handling

The analytic dataset contained deidentified survey responses only. Data were stored in secure study files and accessed by study personnel for analysis. Derived variables used in the manuscript were generated directly from the survey export using the coding rules described above, so that descriptive estimates and univariate models can be reproduced from the raw response file.

### S2. Descriptive statistics

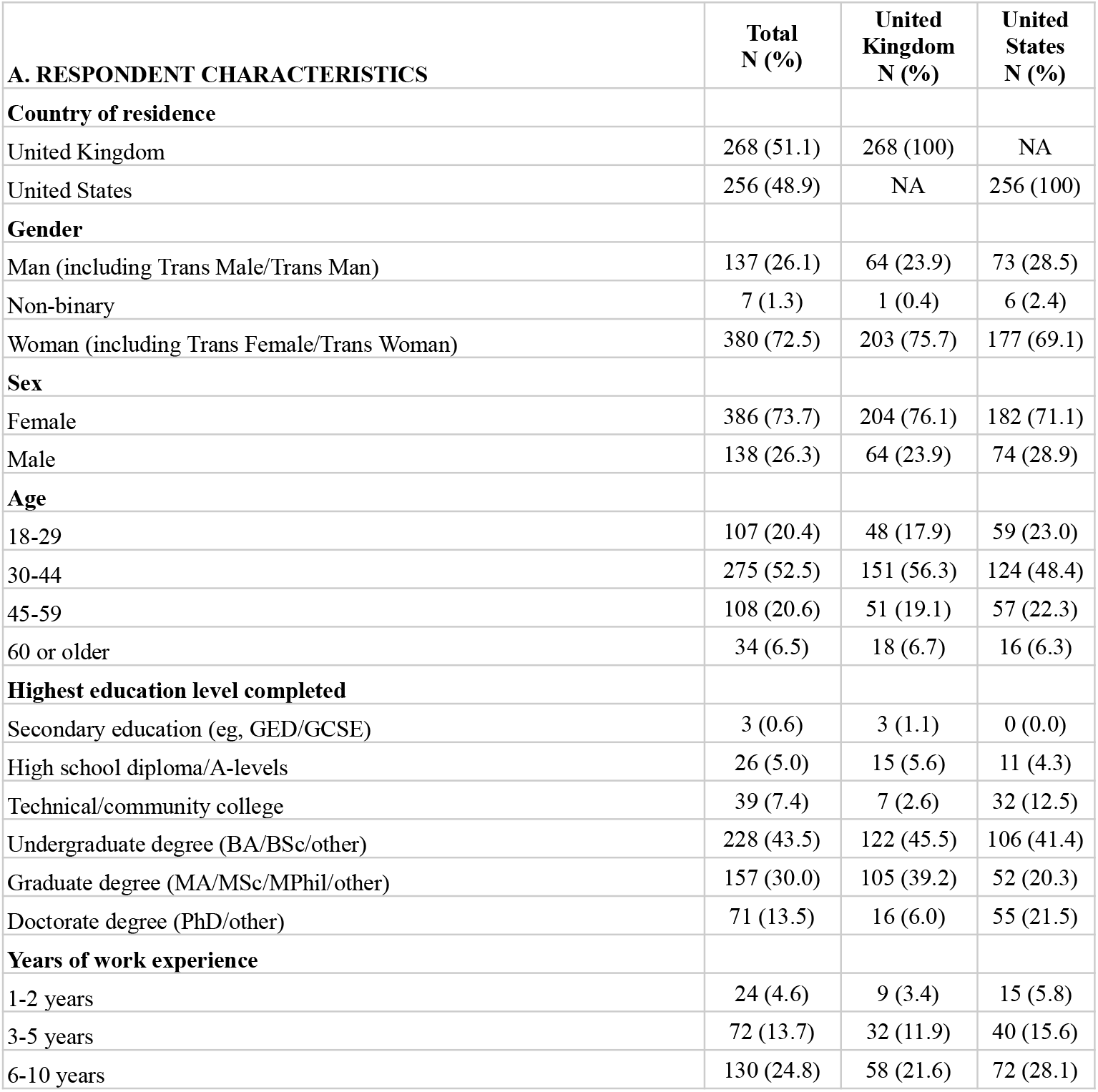
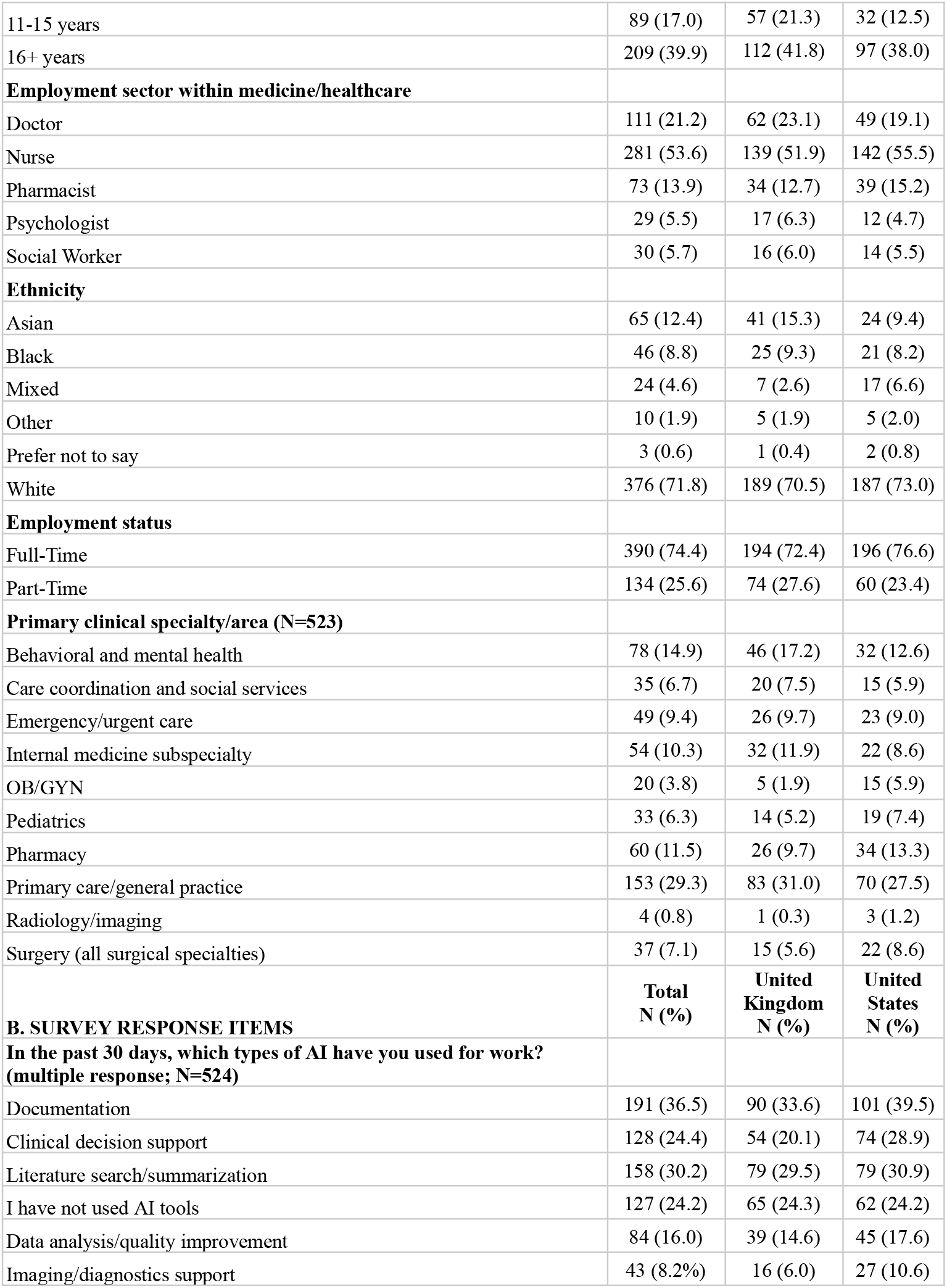
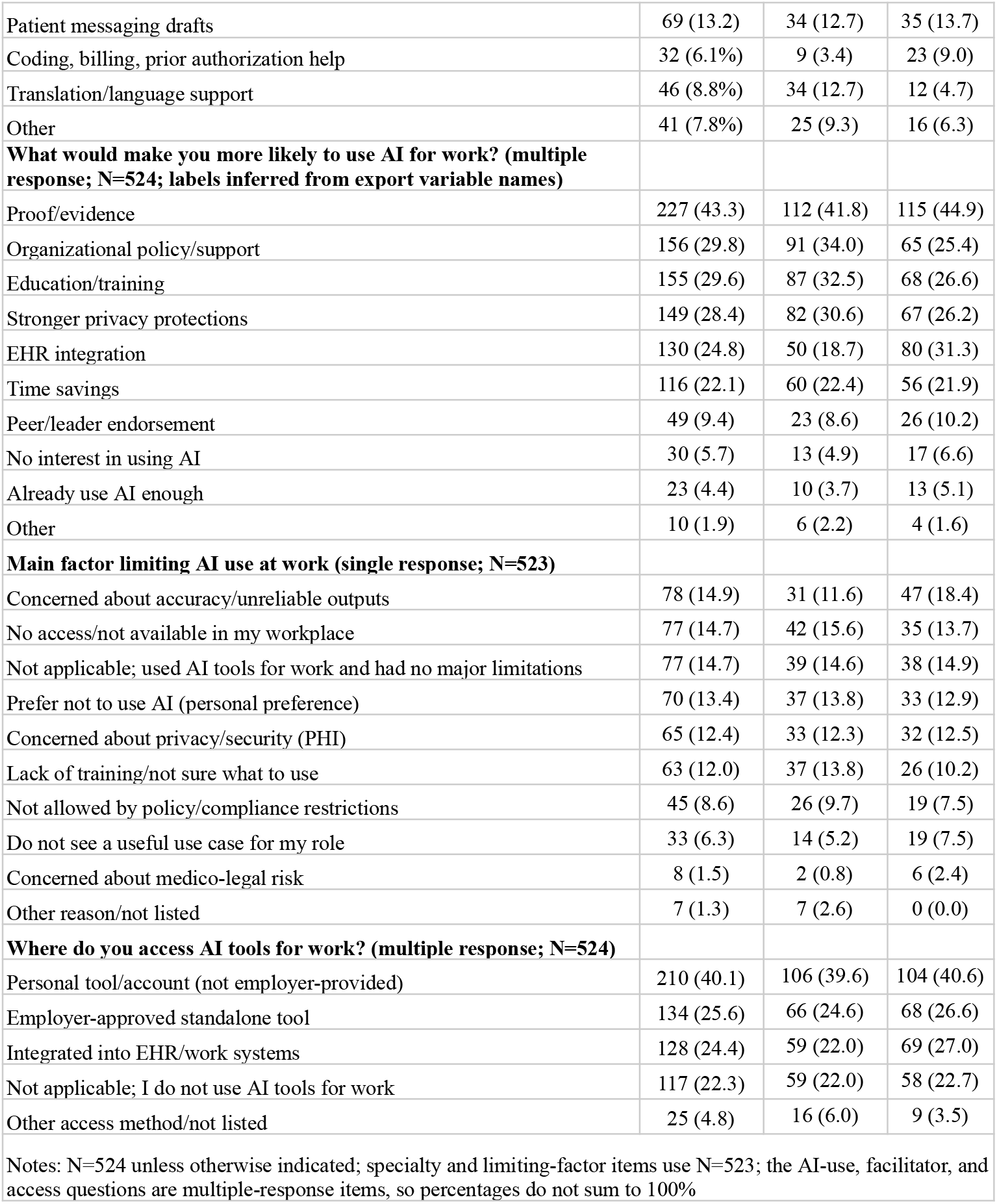

### S3. Univariate Linear Regression

S3. Univariate Linear Regression: Predictors of AI Usage Breadth Score (N=523-524)
|  | Univariate B-coefficient (95% CI) | p-value |
| --- | --- | --- |
| Gender |  |  |
| Male | Reference |  |
| Non-binary | 0.58 (-0.47, 1.62) | 0.28 |
| Female | -0.28 (-0.55, -0.01) | 0.04 |
| Education |  |  |
| High school diploma | Reference |  |
| Secondary education | 0.08 (-1.52, 1.69) | 0.93 |
| Technical/community college | 0.38 (-0.29, 1.06) | 0.26 |
| Undergraduate degree | 0.37 (-0.17, 0.93) | 0.18 |
| Graduate degree | 0.85 (0.29, 1.40) | <0.01 |
| Doctorate degree | 1.03 (0.43, 1.64) | <0.01 |
| Years of work experience |  |  |
| 1-2 years | Reference |  |
| 3-5 years | 0.07 (-0.57, 0.70) | 0.83 |
| 6-10 years | 0.45 (-0.15, 1.05) | 0.14 |
| 11-15 years | 0.38 (-0.24, 1.00) | 0.23 |
| 16+ years | 0.50 (-0.07, 1.08) | 0.09 |
| Age | 0.01 (-0.01, 0.02) | 0.15 |
| Male sex | 0.28 (0.01, 0.55) | 0.04 |
| Role |  |  |
| Doctor | Reference |  |
| Nurse | -0.63 (-0.92, -0.33) | <0.01 |
| Other | -0.61 (-0.94, -0.26) | <0.01 |
| Race |  |  |
| Asian | Reference |  |
| Black | 0.69 (0.17, 1.21) | 0.01 |
| Mixed | 0.08 (-0.56, 0.72) | 0.81 |
| Other/not answer | 0.08 (-0.74, 0.89) | 0.85 |
| White | -0.02 (-0.38, 0.34) | 0.90 |
| Country of Residence |  |  |
| United Kingdom | Reference |  |
| United States | 0.19 (-0.04, 0.42) | 0.11 |
| English as primary language | -0.05 (-0.43, 0.33) | 0.81 |
| Full time employment | 0.37 (0.10, 0.64) | 0.01 |
| Specialty |  |  |
| Primary care/general practice | Reference |  |
| Behavioral Health | -0.31 (-0.68, 0.06) | 0.10 |
| Care Coordination/Social Services | 0.43 (-0.12, 0.97) | 0.12 |
| Emergency Medicine | 0.49 (0.01, 0.98) | 0.05 |
| Internal Medicine | 0.28 (-0.19, 0.76) | 0.24 |
| Obstetrics/gynecology | 0.10 (-0.57, 0.78) | 0.76 |
| Pediatrics | -0.41 (-0.97, 0.15) | 0.15 |
| Pharmacy | 0.01 (-0.46, 0.47) | 0.98 |
| Radiology | -0.35 (-1.72, 1.03) | 0.62 |
| Surgery | 0.01 (-0.53, 0.54) | 0.99 |

Predictors of AI Usage Breadth Score (N=523-524)

AI usage score = sum of AI tool types used in the past 30 days (range 0-9, excluding “not used” response option). Each row represents a separate univariate regression model; N ranged from 523 to 524 because of missing data for some covariates. CI, confidence interval.

